# Clinical Relevance and Applicability of the 2022 World Health Organization Classification of Childhood B Lymphoblastic Leukemia in the Context of MRD-Directed Therapy

**DOI:** 10.1101/2024.10.16.24315431

**Authors:** Sweta Rajpal, Gaurav Chatterjee, Prasanna Bhanshe, Vishram Terse, Swapnali Joshi, Shruti Chaudhary, Dhanalaxmi Shetty, Purvi Mohanty, Chetan Dhamne, Prashant Tembhare, Shyam Srinivasan, Akanksha Chichra, Nirmalya Roy Malik, Shripad Banavali, Sumeet Gujral, Gaurav Narula, PG Subramanian, Nikhil Patkar

**Affiliations:** Department of Haematopathology, ACTREC, Tata Memorial Centre, Navi Mumbai, India; Homi Bhabha National Institute (HBNI), Mumbai, India; Pediatric Haematolymphoid Disease Management Group, Tata Memorial Centre, Mumbai, India; Dept of Cytogenetics, ACTREC, Tata Memorial Centre, Navi Mumbai, India

**Keywords:** B Cell Precursor ALL, RNA sequencing, MRD Directed Therapy

## Abstract

WHO5-2022 classification of B-lymphoblastic leukemia (B-ALL) incorporates several novel entities requiring high-throughput sequencing for their accurate characterization. The clinical relevance of this classification in the context of contemporary MRD-directed therapy is unclear. We analyzed 533 pediatric B-ALL uniformly treated with ICiCLe-ALL-14 protocol as defined by WHO2016 and reclassified them as per WHO5-2022 using targeted sequencing, FISH, and cytogenetics. Subtype-defining genetic abnormalities were identified in 81.2% of the cohort as per the WHO5 classification. Among the new subtypes, *PAX5*^alt^, *MEF2D*-r, and *BCR::ABL1*-like(ABL-class) were associated with an inferior 2-year event-free survival (EFS) of 39.1% (*p*<0.0001), 53.8% (*p*=0.024) and 60.6% (*p*=0.043), respectively. We developed a 3-tier genetic risk stratification model incorporating 15 genetic subtypes and the *IKZF1* deletion. Children with standard, intermediate, and high genetic risk demonstrated 2-year EFS of 92.6%, 71.0%, and 50.7% (p<0.0001), and 2-year overall survival of 94.3%, 81.9%, and 71.6% (p<0.0001), respectively. Genetic risk further identified heterogeneous outcomes among ICiCLe risk groups (p<0.0001). Standard genetic risk was associated with superior OS and EFS irrespective of MRD status. We demonstrate the applicability of the WHO5 classification in routine practice and create a general framework for incorporating the WHO5 classification in risk-adapted therapy for childhood B-ALL.

## Introduction

In the last few decades, successful adoption of risk-directed therapy has resulted in remarkably improved pediatric B-lymphoblastic leukemia/lymphoma (B-ALL) outcomes. Assessment of therapeutic response by detecting and quantitating measurable residual disease (MRD) forms the cornerstone of contemporary risk-adapted protocols.

The underlying biology of the B-ALL subtype, which includes abnormalities of chromosome number such as high hyperdiploid as well as chimeric gene fusions such as *ETV6::RUNX1*, are clinically relevant even in the context of MRD-directed therapies.^1^ Contemporary protocols have thus adopted an integrated approach incorporating both baseline genetics and MRD as an optimal strategy for risk-directed therapy.^1,2^

In the last decade, we have significantly advanced our understanding of B-ALL biology due to progress in genome sequencing technologies such as whole genome and transcriptome sequencing. These seminal studies have led to the identification of novel biological subgroups of B-ALL.^3,4^ The WHO5 and ICC classifications have incorporated several of these new subtypes, such as *TCF3::HLF1*, *PAX5* alteration (*PAX5*^alt^), *PAX5* P80R, *DUX4-*rearrangement (*DUX4-*r)*, MEF2D-*r*, ZNF384*-r*, NUTM1*-r, *ETV6::RUNX1*-like, *IG::MYC*.^5,6^ Recently, Jeha and colleagues have shown the clinical significance of the new subtypes of B-ALL in the context of the MRD-directed St. Jude Total Therapy Study 16 protocol. ^7^ It is important to validate and incorporate these genomics data into clinical practice for prognostication and targeted therapy. ^7,8^

A major challenge in detecting the WHO5-recognised subtypes of B-ALL in routine practice is the heterogenous biology of B-ALL, which requires integration of copy number alterations, fusions involving multiple genes with various partners, and somatic mutations to comprehensively detect all subtypes. For diagnosis of these new entities, legacy workflows like fluorescent in situ hybridization (FISH), multiplexed PCR, and multiplex ligation-dependent probe amplification (MLPA) require multiple rounds of reflexive testing that would result in time delay. Furthermore, PCR-based tests require knowledge of both partners involved in the fusion, which is not always feasible, especially with the novel described entities.

Whole transcriptome sequencing (WTS) has been proposed as a one-stop solution for this problem; however, the clinical translation of WTS in practice is challenging due to the resource-intensive nature and turn-around time of WTS, especially if RNA-sequencing results are to impact therapeutic decisions. We have previously validated NARASIMHA, a lab-developed, cost-effective modular targeted RNA-sequencing assay to detect chimeric gene fusions with knowledge of only one partner gene involved in the fusion (for, e.g., *ABL1*). We hypothesized that a combination of targeted DNA and RNA sequencing, with an initial screen based on legacy techniques, could detect all the entities described in the WHO5 classification of B-ALL.

Here, we present the clinical utility of this approach in detecting WHO5 subtypes in a large, uniformly treated cohort of pediatric B-ALL. We further describe the clinical relevance of such an approach in predicting outcomes in the context of MRD-directed therapy in childhood B-ALL.

## Methods

### Diagnostic Evaluation of B-ALL

#### Diagnosis

A total of 533 newly diagnosed pediatric B-ALL, aged 1-15 years, enrolled in the ICiCLe-ALL-14 trial (Clinical Trials Registry-India number, CTRI/2015/12/006,434), from whom peripheral blood or bone marrow samples were received for molecular testing between January 2019 to March 2022, were included in this study (overview in Supplementary Figure 1). B-ALL diagnosis was made using a 10-12 color, 5-tube multi-color flow cytometry (MFC) panel as per WHO2016 criteria, as described elsewhere.^9^ Also, ploidy status was determined using the FxCycle Violet-dye-based MFC method.^10^ Informed consent was taken from the legal gurdians while enrolling in the ICiCLe-ALL-14 trial (Clinical Trials Registry-India number, CTRI/2015/12/006,434) for participation and data sharing including publication. Institutional Ethics Committee of ACTREC, Tata Memorial Centre gave ethical approval for this work (IEC III – 900924).

#### Cytogenetics Work-up

FISH was performed using locus-specific identifier (LSI) probes for *BCR::ABL1* (Zytovision, Bremerhaven, Germany), *TCF3::PBX1* (CytoTest, Rockville, USA), *ETV6::RUNX1*, and *KMT2A-r* (Abbott, Green Oaks, USA). Hyperdiploid B-ALL was diagnosed using centromeric FISH probes for chromosomes 4, 10, and 17 (Zytovision, Bremerhaven, Germany). SureFISH 7p12.2 IKZF1 (Agilent Dako, USA) probe was used to detect partial/whole *IKZF1* gene deletions. Ploidy analysis was also performed on Giemsa-stained metaphases obtained from overnight cultured BMA/peripheral blood cells to determine the modal chromosome number. Analysis was carried out under a fluorescence microscope (Olympus BX61) using GenASIs software (Applied Spectral Imaging, Israel), and 200 intact interphase nuclei were evaluated by two independent co-observers as previously described.^11^

#### Polymerase Chain Reaction based screening

Common translocations (*BCR::ABL1* (E13/14A2 and E1A2), *ETV6::RUNX1*, *TCF3::PBX1* and *KMT2A::AFF1)* were screened using real-time PCR. *IKZF1* deletions were detected using fragment length analysis.^12,13^

#### Next Generation Sequencing

Targeted RNA sequencing was performed in 392 patients (73.5%) who were negative for common translocations (Supplementary Figure 1, Supplementary Table 1). Patients with negative RNA sequencing (n=124) were subjected to targeted DNA sequencing. The details of these assays and bioinformatics analysis can be seen in supplementary data.

#### Risk stratification and treatment protocol

Patients were stratified according to the ICiCLe-ALL-14 risk stratification detailed in the supplementary data. These patients were treated according to the ICiCLe-ALL-14 risk-adapted protocol.^14^ Briefly, patients underwent four intensive treatment blocks (induction, consolidation, interim maintenance, delayed intensification) followed by 96 weeks of maintenance, varying treatment intensity according to risk group. The first randomization explored the toxicity impact of a shorter induction schedule of prednisolone (3 vs 5 weeks) in young, non-high-risk B-ALL patients. The second randomization assessed the survival benefits of replacing doxorubicin with mitoxantrone during delayed intensification across all patients (Supplementary Figure 2). Only fifteen patients underwent an allogeneic Hematopoietic stem cell transplant. Post-induction (PI) and post-consolidation (PC) MRD assessments were performed using our previously validated highly sensitive MFC assay.^15^

#### Classification according to WHO 2016 and WHO 2022

The pediatric B-ALL cases were assigned to defined genetic categories as per the WHO 2016 (revised 4th edition), and WHO 2022 (5th edition) Classification of Tumours of Haematopoietic and Lymphoid Tissues integrating DNA ploidy by MFC, FISH, chromosomal counting, screening PCR, NARASIMHA and targeted DNA sequencing.^16^

#### Clinical Endpoints and Statistical Analysis

Overall survival (OS) was calculated from the diagnosis date to either the last follow-up date or death with censoring at the last contact. Event-free survival (EFS) was defined from diagnosis to events like death, relapse, or refractory status, also censored at the last contact. Chi-square or Fisher’s exact test (two-sided) in MedCalc (version 14.8.1) assessed the association between categorical variables such as MRD positivity and genetic subgroups. The impact of genetic subgroups on EFS and OS was analyzed using Kaplan-Meier curves, generated on Python3 (version 3.12.0), and compared using the log-rank test. We stratified genetic risk into standard (SGR), intermediate (IGR), and high risk (HGR) based on genetic predictors of outcome and published literature for rare variants. This stratification was correlated with PI MRD and ICiCLe risk groups. Kaplan-Meier curves for these subgroups were produced using MedCalc (version 14.8.1). Multivariate analysis was performed using the Cox proportional-hazards regression analysis.

## Results

### Clinical, laboratory, and risk stratification characteristics of patients enrolled

Our final analysis included 533 children with B-ALL with a male-to-female ratio of 1.99:1 and a median age of 5 years. According to the NCI risk stratification, most patients were classified as standard risk (SR, 67.2%). However, with the final ICiCLe risk stratification, the majority (57.4%) were categorized as high risk (HiR), followed by SR (22.9%) and intermediate risk (IR, 19.7%). Nearly one-third of patients (32.2%, out of 519 assessed) were detected to harbor measurable residual disease (defined as MRD ≥0.01%) at the PI timepoint. Similarly, 9.8% (out of 244 assessed) were positive at the PC timepoint. A summary of clinical, laboratory, and genetic characteristics, as well as MRD responses, can be seen in Table 1.

**Table 1.** Demographic and Laboratory Characteristics, WHO 2016 Classification, MRD characteristics and Risk Stratification of B-ALL Patients.

|  |  |
| --- | --- |
| **Demographics** |  |
| Total number of patients accrued | 533 |
| Age (years) | Range:- 1-15, Median:- 5 |
| Gender | Male: Female- 1.99:1 |
| **Laboratory characteristics** |  |
| TLC at presentation |  |
| 1. $\geq 50 \times 10^9/L$ | 81 (15.2%) |
| 2. $< 50 \times 10^9/L$ | 452 (84.8%) |
| **WHO 2016 Genetic categories** |  |
| 1. Hyperdiploidy | 182 (34.1%) |
| 2. *ETV6::RUNX1* | 68 (12.8%) |
| 3. *TCF3::PBX1* | 42 (7.9%) |
| 4. *BCR::ABL1* | 29 (5.4%) |
| 5. *BCR::ABL1* like | 24 (4.5%) |
| 6. *KMT2A* rearranged | 19 (3.6%) |
| 7. Hypodiploidy | 6 (1.1%) |
| 8. iAMP21 | 3 (0.6%) |
| 9. NOS | 160 (30.0%) |
| **NCI Risk** |  |
| SR | 358 (67.2%) |
| HR | 175 (32.8%) |
| **ICiCLe Initial Risk (n=533)** |  |
| SR | 205 (38.4%) |
| IR | 148 (27.8%) |
| HR | 180 (33.8%) |
| **ICiCLe Risk (n=519)** |  |
| SR | 119 (22.9%) |
| IR | 102 (19.7%) |
| HR | 298 (57.4%) |
| **Post-induction MRD (n=519)** |  |
| Positive | 167 (32.2%), Median- 0.14, Range- 0.01-88 |
| Negative | 352 (67.8%) |
| **Post-consolidation MRD (n=244)** |  |
| Positive | 24 (9.8%), Median- 0.2, Range- 0.01-86 |
| Negative | 220 (90.2%) |

### Characterization of Genetic Subgroups and their Correlations

Through the integration of conventional PCR, targeted RNA sequencing, cytogenetics, and immunophenotyping, we identified 13 distinct genetic subgroups within our cohort (Table 2). *PAX5 ^alt^*(24, 4.5%), *BCR::*ABL1-like (23, 4.3%), *DUX4*-r (12, 2.3%), *MEF2D*-r (13, 2.4%), *ZNF384*-r (3, 0.6%) and *TCF3::HLF* (4, 0.7%) subgroups of B-ALL were exclusively identified using RNA-sequencing (Figure 1). Among the *BCR::ABL1*-like category, almost equal division was seen among the *BCR::ABL1*-like: ABL-class (n-11) and JAK-STAT activated (n-13) classes. Among the *BCR::ABL1*-like: ABL-class, the 3’ partner were *PDGFRB* (*EBF1*; n=5), *ABL1* (n=4; one each of *ETV6*, *NUP214*, *RANBP2*, *FOXP1*), *ABL2* (*RCSD1*) and *CSF1R* (*LBD1*) and those in JAK-STAT activated class were *CRLF2* (n=9; *P2RY8;* n=8 and one *IGH*), *JAK2* (n=4; comprising *PAX5*; n=3 and one *EBF1*). For *PAX5*^alt^ category, the 3’ fusion partners were *AUTS2* (n=5), succeeded by *MBNL1* (n=3), *ETV6* (n=2), *IGH* (n=2), *ZNF521* (n=2) and one each with *CBFA2T3*, *FOXP1*, *GSE1*, *ANKRD12, CA10,* and *ESSRB*. For *MEF2D*-r, the fusion partners were *BCL9* (n=8), *HNRNPUL* (n=3), *DASAP1* (n=1) and *SF1* (n=1). For *ZNF384*-r cases, the fusion partners were *TCF3* (n=2) and *EP300* (n=1).

**Figure 1.**
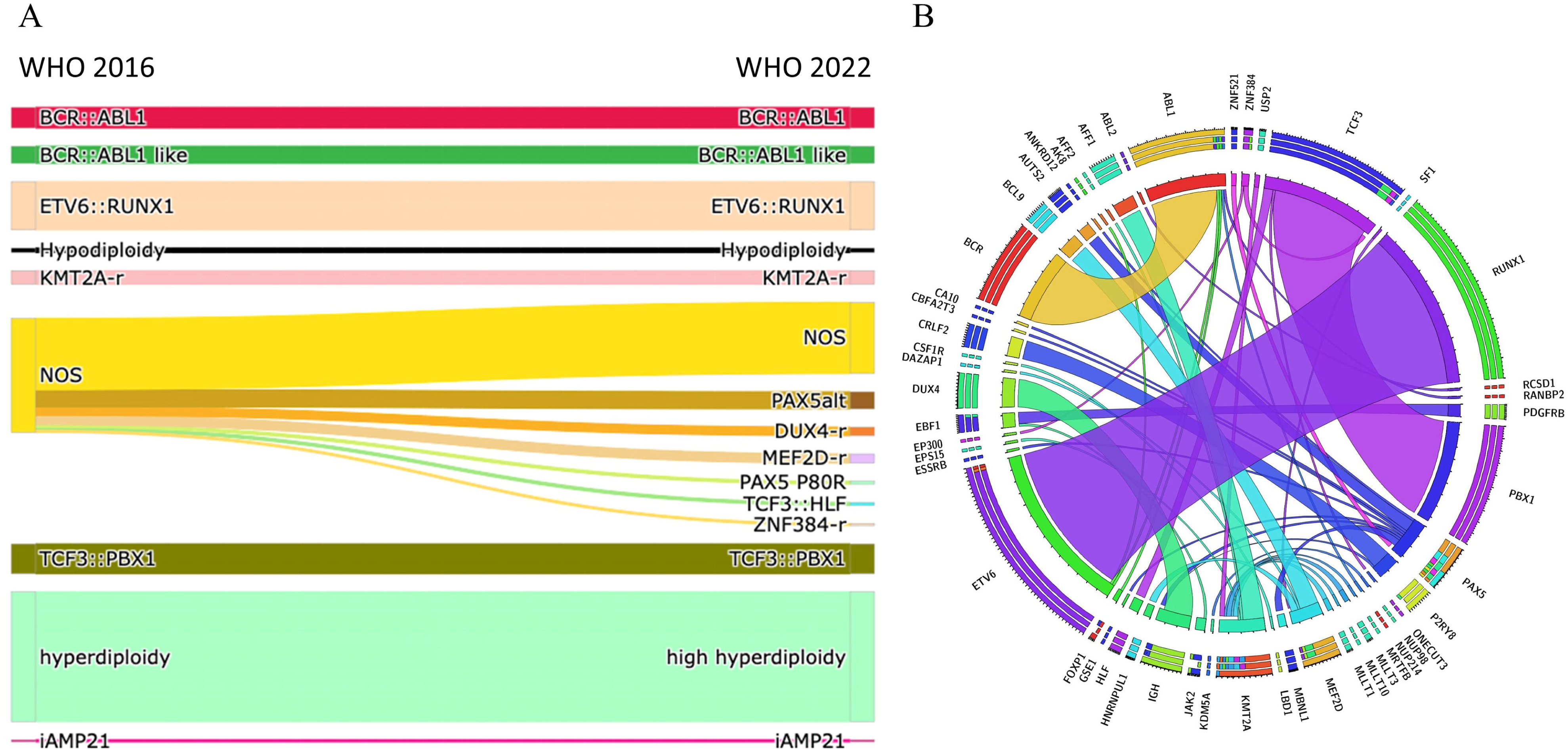
(a) Sankey Diagram Illustrating the Reclassification of B-ALL Cases from WHO 2016 to WHO 2022 B-ALL Classification (b) Circos diagram showing fusions identified in pediatric B-ALL.

**Table 2.** Age and Sex Distribution, WBC Counts, and MRD characteristics across Genetic Subgroups. * p-values are not provided for genetic subgroups with fewer than 10 cases.

| Genetic category | N (%) | Median age in years (range), p-value* | Male n (%), p-value* | Median WBC (range) at diagnosis (X 10<sup>9</sup>/L), p-value* | Percentage of PI MRD positive cases (median, range), p-value* |
| --- | --- | --- | --- | --- | --- |
| **All categories** | **533** | **5 (1-15)** | **355 (66.5)** | **6.37 (0.23-692.3)** | **32.2 (0.14, 0.01- 88)** |
| High hyperdiploidy | 182 (34.1) | 4 (1 – 13), < 0.001 | 125 (67.2), 1.000 | 4.63 (0.61–253.0), < 0.001 | 30.3 (0.01, 0.01- 11), 0.38 |
| *ETV6::RUNX1* | 68 (12.8) | 4 (1 – 15), 0.003 | 44 (66.7), 1.000 | 4.85 (0.23–90.1), 0.064 | 24.2 (0.02, 0.01-5.8), 0.10 |
| *TCF3::PBX1* | 42 (7.9) | 6(2-14), 0.154 | 24 (57.1), 0.177 | 14.185 (1.22–281.0), 0.003 | 26.2 (0.03, 0.01-3.60), 0.55 |
| *BCR::ABL1* | 29 (5.4) | 10 (2 – 14), <0.001 | 22 (75.9), 0.318 | 61.71 (1.11–559.4), <0.01 | 48.3 (0.48, 0.01-23.50), 0.01 |
| *BCR::ABL1* like | 24 (4.5) | 11.5 (3-15), 0.001 | 17 (70.8), 0.825 | 33.525 (0.93–295.1), 0.005 | 70.8 (1.6, 0.01-62), 0.000033 |
| *PAX<sup>alt</sup>* | 24 (4.5) | 4 (1-14), 0.354 | 16 (69.6), 0.825 | 8.15 (0.59–208.5), 0.637 | 23.8 (0.04, 0.01-1.1), 0.46 |
| *KMT2A-r* | 19 (3.6) | 3 (0-14), 0.001 | 12 (63.2), 0.806 | 19.54 (1.37–424.71), 0.055 | 26.3 (0.26, 0.01-50), 0.76 |
| *MEF2D-r* | 13 (2.4) | 9 (3-14), 0.004 | 5 (38.5), 0.038 | 11.15 (1.31–119.11), 0.252 | 30.8 (0.05, 0.01- 0.24), 1.00 |
| *DUX4-r* | 12 (2.3) | 6 (3-14), 0.167 | 9 (75.0), 0.759 | 5.14 (1.56–27.73), 0.358 | 33.3 (0.03, 0.01-3.5), 1.00 |
| Hypodiploidy | 6 (1.1) | 11 (4-13) | 4 (66.7) | 3.27 (1.34–155.1) | 33.3 (0.01, 0.01-4.3) |
| *PAX5 P80R* | 4 (0.7) | 12 (8-14) | 4 (100) | 55.87 (1.99-86.8) | 0 |
| *TCF3::HLF* | 4 (0.7) | 8 (4-14) | 3 (75) | 6.335 (1.32–9.74) | 75.0 (1.25, 0.01- 6.5) |
| *ZNF384-r* | 3 (0.6) | 5 (4-16) | 1 (33.3) | 14.33 (4.37–22.32) | 66.7 (6.20, 0.32-12.09) |
| iAMP21 | 3 (0.6) | 6 (2-10) | 2 (66.7) | 3.04 (2.94-5.4) | 0, 0.56 |
| NOS | 100 (18.8) | 7 (1 – 14), < 0.001 | 70 (70.0), 0.481 | 8.07 (0.29-692.3), 0.147 | 33.0 (0.05, 0.01-71.7), 1.00 |

Targeted DNA sequencing further helped identify additional *PAX5*^alt^ (n=4) and the *PAX5* P80R group (n=4). Using this approach, we could identify a genomic abnormality in 81.2% of all cases when compared to legacy approaches (61.7%) Patients harboring *BCR::ABL1* (median age-10 years; p<0.001)*, BCR::ABL1*-like (median age-11.5 years, *p*=0.001), and *MEF2D*-r (median age-9 years; *p*=0.004) were significantly older compared to the entire cohort (median age-5 years; range-1 to 15 years). *BCR::ABL1* (median WBC-61.7 x 10^9^/L; *p*=0.003) and *BCR::ABL1* like (median WBC-33.5 x 10^9^/L; *p*=0.005) showed higher WBC counts as compared to the entire cohort (median WBC-6.4 x 10^9^/L; range-0.2 to 692.3 x 10^9^/L). Additionally, the *BCR::ABL1* (48.3% MRD-positive; median MRD: 0.48; *p*=0.02) and *BCR::ABL1*-like (70.8% MRD-positive; median MRD: 1.6; *p*<0.0001) groups exhibited higher PI MRD positivity as compared to the rest (32.3% PI MRD-positive; median PI MRD: 0.14%). The MRD characteristics of each sub-group are detailed in supplementary data and highlighted in Supplementary Figure 3.

*IKZF1* deletion was identified in 69 (15.1%) of the 458 patients tested and showed a high prevalence in the *BCR::ABL1* (56.5%) and *BCR::ABL1*-like (55.6%) genetic subgroups. Additionally, *IKZF1* deletions were detected across various other subgroups, including NOS, high hyperdiploidy, *PAX5*^alt^, *DUX4*-r, *KMT2A*-r, hypodiploidy, *ETV6::RUNX1*, and iAMP21 (Supplementary Figure 4).

### Re-stratification from the WHO 2016 to WHO 2022 classification

There was no change in the classification of 373 patients (*ETV6::RUNX1,* High hyperdiploidy, *BCR::ABL1, TCF3::PBX1, BCR::ABL1* like *KMT2A-r*, hypodiploidy, iAMP21) when they were classified as per the WHO 2022 compared to the WHO 2016. Among WHO 2016-defined NOS subgroup (n=160, 30% of the entire cohort), 60 (37.5%) children were reclassified into newly recognized subtypes such as *PAX5^alt^* (n=24, 15%), *DUX4*-r (n=12, 7.5%), *MEF2D*-r (n=13, 8.1%), *PAX5* P80R (n=4, 2.5%), *TCF3::HLF* (n=4, 2.5%), and *ZNF384-r* (n=3, 1.9%) as seen in Figure 1A.

### Patient Outcomes

The 2-year EFS and OS of the entire cohort were 75.8% and 85.1%, respectively (median EFS and OS not reached). The median follow-up of the cohort was 26.8 months. The presence of PI MRD predicted a significantly inferior EFS [HR-2.0; 95% CI-1.42 to 2.89; (*p*<0.0001)] and OS [HR-1.7; 95% CI-1.08 to 2.64; (*p*=0.01)]. Similarly, PC MRD positivity was associated with markedly worse EFS [HR-22.0; 95% CI-4.37 to 110.93; (*p*<0.0001)] and OS [HR-2.4; 95% CI-0.78 to 7.70; (*p*=0.02)] (Supplementary Figure 5). The impact of ICiCLe risk stratification on patient outcomes is detailed in the supplementary data (Supplementary Figure 6).

### Outcomes for individual genetic subgroups

On evaluating the impact of genetic subgroups, the following were associated with inferior EFS: *PAX5*^alt^ (HR-2.8; 95% CI-1.18 to 6.68; *p*=0.0001), *BCR::ABL1* (HR-2.5; 95% CI-1.17 to 5.51; *p*=0.0002), *MEF2D*-r (HR-2.3; 95% CI-0.75 to 7.20; *p*=0.024), and *BCR::ABL1*-like with ABL-class fusion (HR-2.3; 95% CI-0.68 to 7.55; *p*=0.043) (Table 3). Subgroups associated with inferior OS were *PAX5*^alt^ (HR-4.3; 95% CI-1.46 to 12.96; *p*<0.0001) and *BCR::ABL1* (HR-2.4; 95% CI-0.98 to 6.11; *p*=0.004). Similarly, *IKZF1* deletion was associated with inferior EFS (HR-3.0; 95% CI-1.72 to 5.07; *p*<0.0001) and OS (HR-1.8; 95% CI-0.95 to 3.51; *p*=0.022). *TCF3::HLF* subgroup also exhibited poor EFS, with three out of four affected children having refractory disease and one experiencing an early relapse; however, statistical analysis was not performed due to the small sample size.

**Table 3.** Event-free and overall survival rates for genetic categories of B-ALL, as classified by WHO 2022, including hazard ratios and p-values. The genetic categories with fewer than 10 cases are not included in the table.

|  | Overall survival |  |  | Event free survival |  |  |
| --- | --- | --- | --- | --- | --- | --- |
|  | 2 years OS | P-value | Hazard ratio (95% Confidence Interval) | 2 years EFS | P-value | Hazard ratio (95% Confidence Interval) |
| *IKZF1* del | 77.5 | 0.022 | 1.8 (0.95 to 3.51) | 46.2 | <0.0001 | 3.0 (1.72 to 5.07) |
| *PAX5*<sup>alt</sup> | 46.6 | <0.0001 | 4.3 (1.46 to 12.96) | 39.1 | 0.0001 | 2.8 (1.18 to 6.68) |
| *BCR::ABL1* | 67.6 | 0.004 | 2.4 (0.98 to 6.11) | 50.6 | 0.0002 | 2.5 (1.17 to 5.51) |
| *MEF2D-r* | 66.1 | 0.114 | 2.2 (0.51 to 9.33) | 53.8 | 0.024 | 2.3 (0.75 to 7.20) |
| *BCR::ABL1* like, ABL-Class | 90.9 | 0.821 | 0.9 (0.23 to 3.11) | 60.6 | 0.043 | 2.3 (0.68 to 7.55) |
| NOS | 84.2 | 0.714 | 1.1 (0.67 to 1.79) | 65.9 | 0.015 | 1.6 (1.03 to 2.33) |
| *TCF3::PBX1* | 77.3 | 0.133 | 1.6 (0.77 to 3.26) | 67.4 | 0.253 | 1.3 (0.76 to 2.36) |
| Hypodiploidy | 83.3 | 0.961 | 1.1 (0.14 to 7.90) | 66.7 | 0.684 | 1.3 (0.27 to 6.64) |
| *KMT2A-r* | 72.0 | 0.214 | 1.8 (0.5469 to 5.6249) | 67.5 | 0.645 | 1.2 (0.50 to 2.95) |
| *BCR::ABL1* like, JAK-STAT | 84.6 | 0.730 | 1.2 (0.35 to 4.33) | 63.5 | 0.98 | 1.0 (0.37 to 2.65) |
| *DUX4-r* | 82.5 | 0.791 | 0.8 (0.23 to 2.97) | 73.3 | 0.58 | 0.7 (0.27 to 1.94) |
| *ETV6::RUNX1* | 100 | 0.027 | 0.4 (0.23 to 0.72) | 98.0 | 0.006 | 0.4 (0.27 to 0.65) |
| High Hyperdiploidy | 92.1 | 0.0006 | 0.4 (0.29 to 0.65) | 88.9 | <0.0001 | 0.4 (0.26 to 0.49) |

Conversely, high hyperdiploidy was associated with superior EFS (HR-0.4; 95% CI-0.26 to 0.49; *p* < 0.0001) and OS (HR-0.4; 95% CI-0.29 to 0.65; *p*=0.0006). *ETV6::RUNX1* also predicted superior EFS (HR-0.4; 95% CI-0.27 to 0.65; *p*=0.006) and OS (HR-0.4; 95% CI-0.23 to 0.72; *p*=0.027) as seen in Table 3.

### Genetic risk assignment

We devised a 3-tier genetic risk model (SGR, IGR, and HGR) incorporating variables found to be significantly predictive of EFS on univariate analysis in the present study. Furthermore, if the number of available cases was <10 in our study, available literature for those categories was considered (for, e.g., *TCF3::HLF*). Genetic categories with a hazard ratio (HR) < 0.75 were classified as SGR, those with an HR between 0.75 and 2 as IGR, and those with an HR > 2 as HGR. The details of this classification can be seen in Figure 3. The 2-year EFS for SGR, IGR, and HGR was 92.6%, 71.2%, and 50.3%, respectively (p<0.0001), and the 2-year OS for SGR, IGR, and HGR were 94.3%, 81.9%, and 71.6%, respectively (p<0.0001; Table 4).

**Figure 2.**
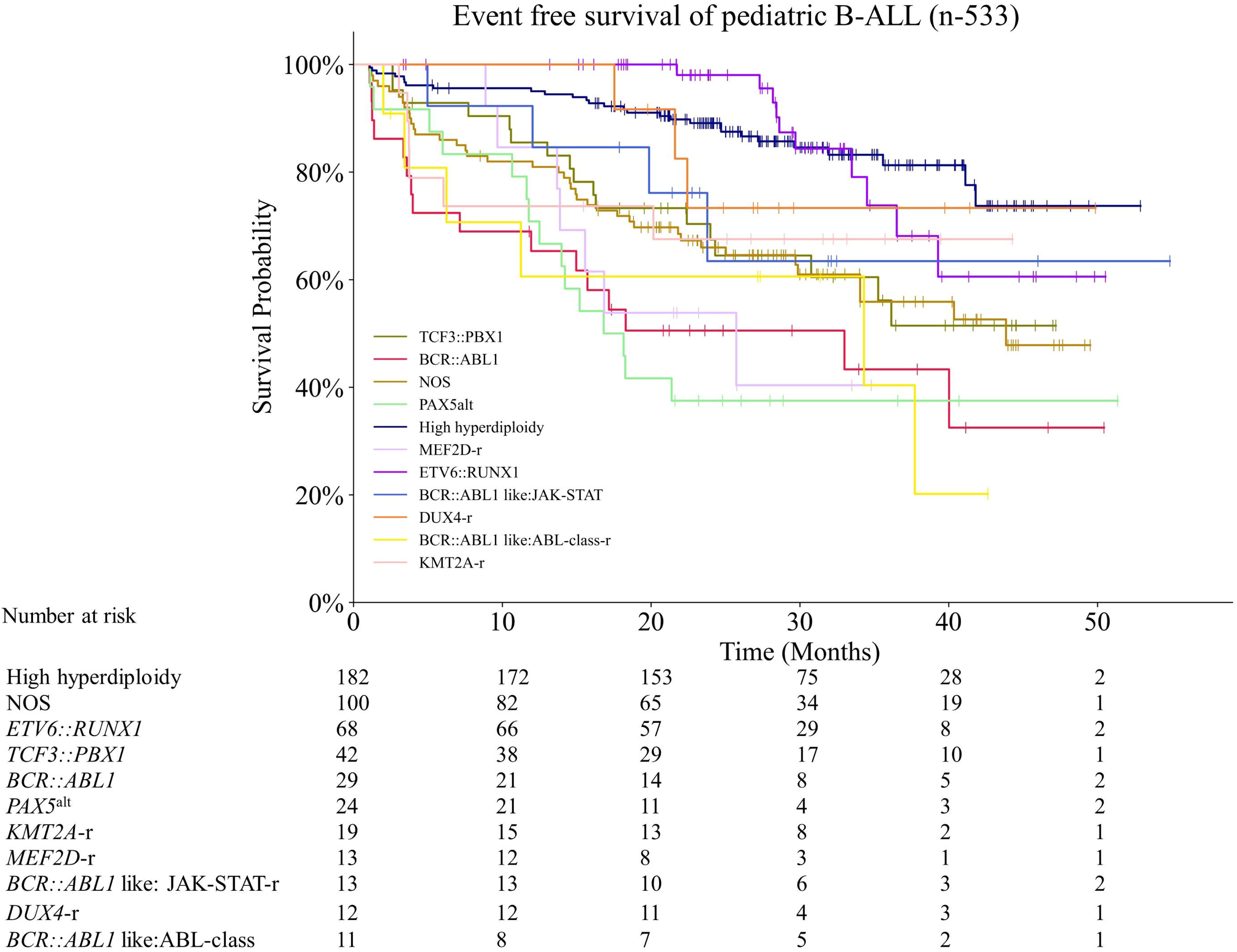
Event-free survival (EFS) of pediatric B-cell acute lymphoblastic leukemia (B-ALL) patients (n=533) categorized by genetic subtypes.

**Figure 3.**
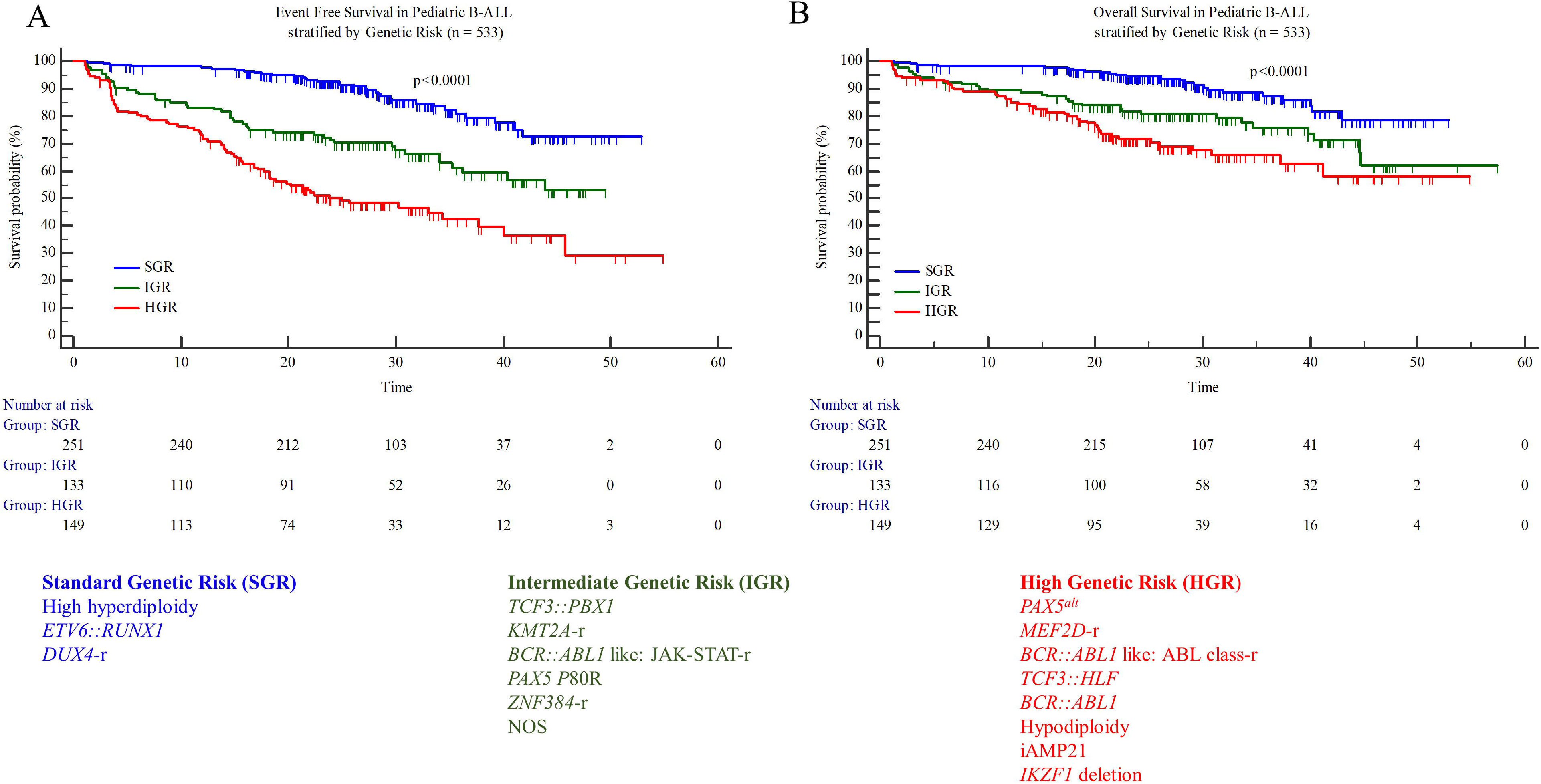
Event-free and overall survival of pediatric B-ALL patients stratified by genetic risk classification into standard (SGR), intermediate (IGR), and high (HGR) genetic risk groups.

**Table 4.**
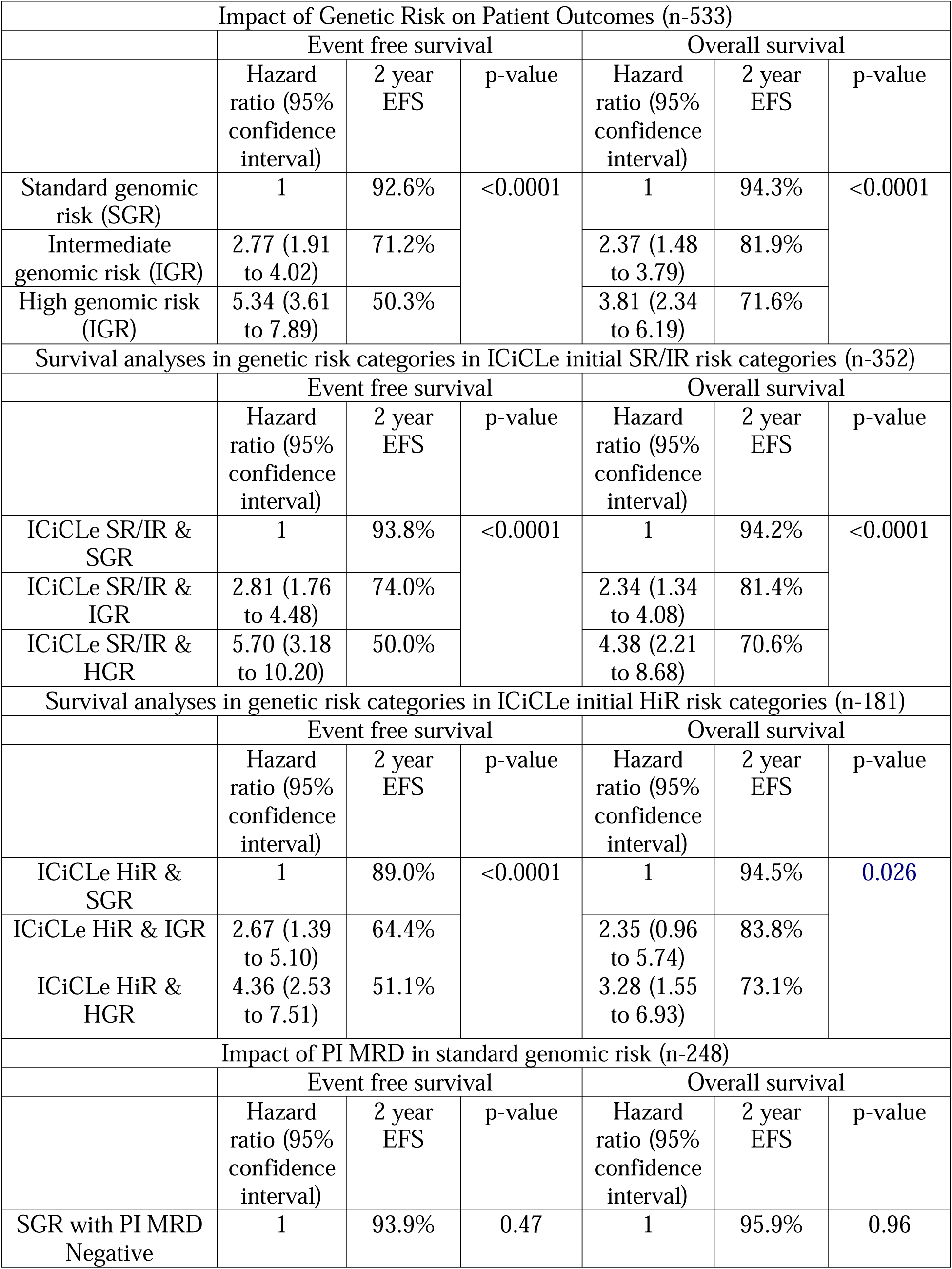
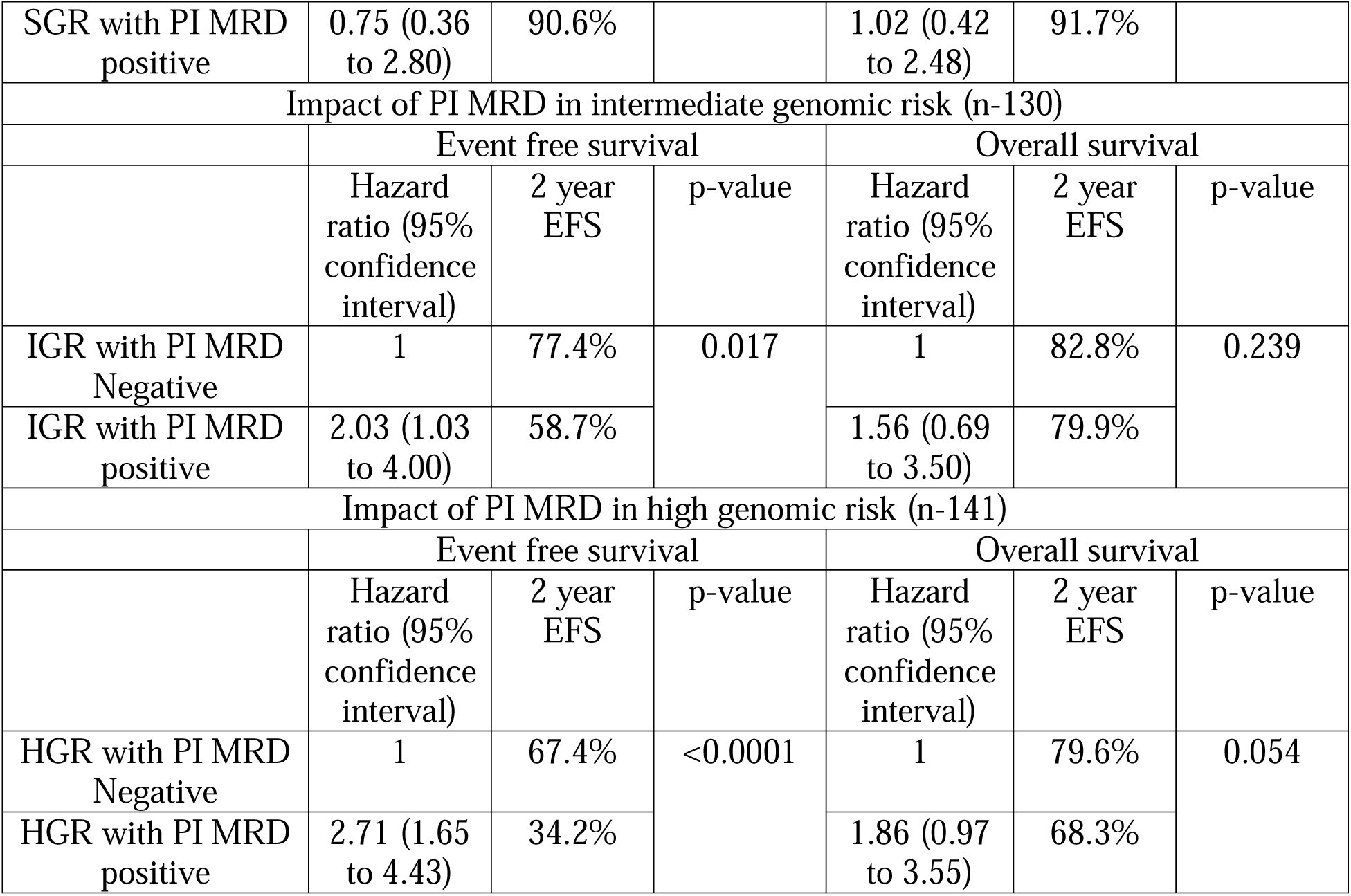
Genetic risk-based survival outcomes and the impact of post-induction measurable residual disease positivity.

| Impact of Genetic Risk on Patient Outcomes (n-533) |  |  |  |  |  |  |
| --- | --- | --- | --- | --- | --- | --- |
|  | Event free survival |  |  | Overall survival |  |  |
|  | Hazard ratio (95% confidence interval) | 2 year EFS | p-value | Hazard ratio (95% confidence interval) | 2 year EFS | p-value |
| Standard genomic risk (SGR) | 1 | 92.6% | <0.0001 | 1 | 94.3% | <0.0001 |
| Intermediate genomic risk (IGR) | 2.77 (1.91 to 4.02) | 71.2% |  | 2.37 (1.48 to 3.79) | 81.9% |  |
| High genomic risk (IGR) | 5.34 (3.61 to 7.89) | 50.3% |  | 3.81 (2.34 to 6.19) | 71.6% |  |
| Survival analyses in genetic risk categories in ICiCLE initial SR/IR risk categories (n-352) |  |  |  |  |  |  |
|  | Event free survival |  |  | Overall survival |  |  |
|  | Hazard ratio (95% confidence interval) | 2 year EFS | p-value | Hazard ratio (95% confidence interval) | 2 year EFS | p-value |
| ICiCLE SR/IR & SGR | 1 | 93.8% | <0.0001 | 1 | 94.2% | <0.0001 |
| ICiCLE SR/IR & IGR | 2.81 (1.76 to 4.48) | 74.0% |  | 2.34 (1.34 to 4.08) | 81.4% |  |
| ICiCLE SR/IR & HGR | 5.70 (3.18 to 10.20) | 50.0% |  | 4.38 (2.21 to 8.68) | 70.6% |  |
| Survival analyses in genetic risk categories in ICiCLE initial HiR risk categories (n-181) |  |  |  |  |  |  |
|  | Event free survival |  |  | Overall survival |  |  |
|  | Hazard ratio (95% confidence interval) | 2 year EFS | p-value | Hazard ratio (95% confidence interval) | 2 year EFS | p-value |
| ICiCLE HiR & SGR | 1 | 89.0% | <0.0001 | 1 | 94.5% | 0.026 |
| ICiCLE HiR & IGR | 2.67 (1.39 to 5.10) | 64.4% |  | 2.35 (0.96 to 5.74) | 83.8% |  |
| ICiCLE HiR & HGR | 4.36 (2.53 to 7.51) | 51.1% |  | 3.28 (1.55 to 6.93) | 73.1% |  |
| Impact of PI MRD in standard genomic risk (n-248) |  |  |  |  |  |  |
|  | Event free survival |  |  | Overall survival |  |  |
|  | Hazard ratio (95% confidence interval) | 2 year EFS | p-value | Hazard ratio (95% confidence interval) | 2 year EFS | p-value |
| SGR with PI MRD Negative | 1 | 93.9% | 0.47 | 1 | 95.9% | 0.96 |
| SGR with PI MRD positive | 0.75 (0.36 to 2.80) | 90.6% |  | 1.02 (0.42 to 2.48) | 91.7% |  |
| Impact of PI MRD in intermediate genomic risk (n-130) |  |  |  |  |  |  |
|  | Event free survival |  |  | Overall survival |  |  |
|  | Hazard ratio (95% confidence interval) | 2 year EFS | p-value | Hazard ratio (95% confidence interval) | 2 year EFS | p-value |
| IGR with PI MRD Negative | 1 | 77.4% | 0.017 | 1 | 82.8% | 0.239 |
| IGR with PI MRD positive | 2.03 (1.03 to 4.00) | 58.7% |  | 1.56 (0.69 to 3.50) | 79.9% |  |
| Impact of PI MRD in high genomic risk (n-141) |  |  |  |  |  |  |
|  | Event free survival |  |  | Overall survival |  |  |
|  | Hazard ratio (95% confidence interval) | 2 year EFS | p-value | Hazard ratio (95% confidence interval) | 2 year EFS | p-value |
| HGR with PI MRD Negative | 1 | 67.4% | <0.0001 | 1 | 79.6% | 0.054 |
| HGR with PI MRD positive | 2.71 (1.65 to 4.43) | 34.2% |  | 1.86 (0.97 to 3.55) | 68.3% |  |

### Genetic Risk Identifies Heterogeneous Outcomes within ICiCLe Risk Groups

Since patients in this cohort were treated with risk-adapted protocol, we further evaluated the effect of genetic risk stratification in risk groups treated with similar intensities. The ICiCLe initial SR and IR were combined for this analysis, while the HiR group was analyzed separately. Genetic risk stratification retained its prognostic significance within both ICiCLe initial SR/IR and HiR groups. The 2-year EFS of SGR, IGR, and HGR within ICiCLe initial SR/IR were 93.8%, 74.0%, and 50.0%, respectively (p<0.0001), and that within ICiCLe initial HiR was 89.2%, 64.4% and 51.1% respectively (p<0.0001) (Figure 4). Similarly, the 2-year OS of SGR, IGR, and HGR within ICiCLe initial SR/IR were 94.2%, 81.4%, and 70.6%, respectively (p<0.0001), and that within ICiCLe initial HiR was 94.5%, 83.8% and 73.1% respectively (*p*=0.026) (Table 4).

**Figure 4.**
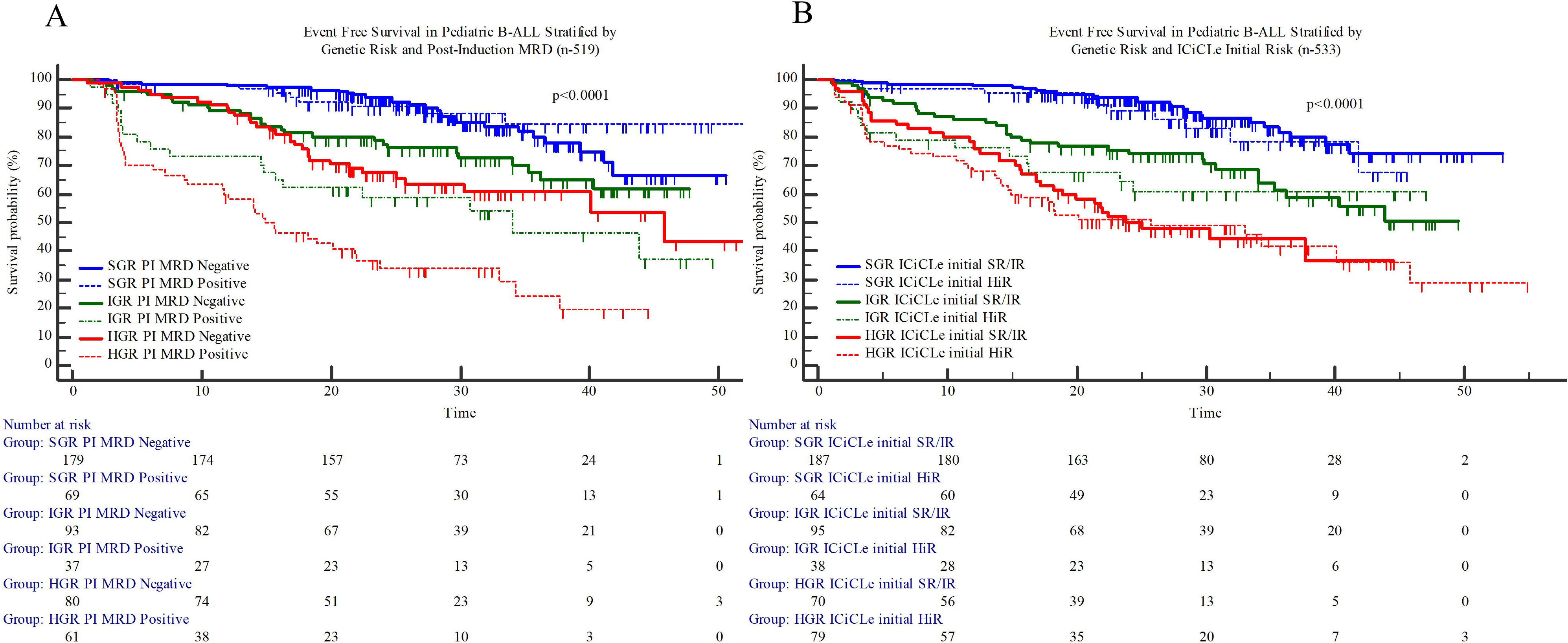
(a) Event free survival in pediatric B-ALL stratified by genetic risk and post-induction MRD revealing similar outcome in the standard genomic risk category irrespective of post-induction measurable residual disease status; (b) Event free survival in pediatric B-ALL stratified by ICiCLe initial risk stratification and genetic risk heterogenous outcomes within the ICiCLe risk groups.

### Genetic Risk Identifies Heterogeneity in Standard Risk B-ALL Irrespective of MRD Status

Patients in the SGR group exhibited superior outcomes regardless of PI MRD status, with 2-year EFS rates of 93.9% for MRD-positive and 90.6% for MRD-negative patients (*p*=0.469), and 2-year OS rates of 95.9% and 93.5%, respectively (*p*=0.954). In the IGR group, OS was similar between MRD-negative (84.2%) and MRD-positive (79.9%) patients (*p*=0.239). In the HGR group, OS was 79.6% for MRD-negative and 68.3% for MRD-positive patients (*p*=0.054). However, MRD status remained a significant predictor of EFS in the IGR and HGR groups, with 2-year EFS rates of 58.7% for MRD-positive versus 77.4% for MRD-negative in the IGR group (*p*=0.017), and 34.2% for MRD-positive versus 67.4% for MRD-negative in the HGR group (p<0.0001) (Table 4, Figure 4).

### Multivariate analysis

On multivariate analysis, genetic risk stratification (HR-2.0; 95% CI-1.62 to 2.45; *p<*0.0001), initial ICiCLe risk (HR-1.3; 95% CI-1.05 to 1.57; *p=*0.015), and PI MRD (HR-2.0; 95% CI-1.42 to 2.74; *p<*0.0001) independently predicted EFS, with genetic risk stratification (HR-1.7; 95% CI-1.29 to 2.17; *p=*0.0001) and PI MRD positivity (HR-1.6; 95% CI-1.03 to 2.38; *p<*0.0001) also independently impacting OS (Supplementary Table 2).

## Discussion

Pioneering recent studies have utilized unbiased broad-spectrum approaches such as WTS/WGS to uncover novel genetic subtypes^4,17^, which have been recognized as distinct entities in the latest WHO5/ICC classification of B-ALL.^16^ Routine clinical application of WTS/WGS assays remains challenging. Herein, we demonstrate that targeted RNA and DNA sequencing, in combination with standard FISH/cytogenetics, is routinely implementable in clinical practice and can detect all of the WHO5-recognized subtypes of B-ALL. We demonstrate that a targeted sequencing approach identifies prognostically relevant subgroups even in the era of contemporary MRD-directed therapies.

Among the recently defined genetic categories, we found frequencies of 4.5% for *BCR::ABL1*-like, 4.5% for *PAX5*^alt^, 2.44% for *MEF2D*-r, 2.25% for *DUX4*-r, and 0.75% each for *PAX5* P80R, and *TCF3::HLF*, and 0.56% for *ZNF384*-r. The proportion of *PAX5*^alt^ (4.5% vs. 7.5%)^4^, and *DUX4*-r (2.25% vs. 4%)^3^ cases in our cohort are lower than that reported in the discovery cohorts. The NARASIMHA assay used in this study focuses only on detecting chimeric gene fusions. Unbiased sequencing approaches such as WTS that include gene expression profiling might have uncovered further cases of *BCR::ABL1*-like and *DUX4*-r. Additionally, we could not detect other specific categories, such as ETV6::RUNX1-like, due to a lack of gene expression profiling. Another factor to consider is that we report results only on a large cohort of children uniformly recruited in the ICiCLe-ALL-14 trial. Interestingly, transcriptomic sequencing findings from the St. Jude Total Therapy Study 16 trial reported similar *BCR::ABL1*-like (3%) and *PAX5*^alt^ (4.85%) cases.^7^ In total, 81.2% of the children analyzed could be assigned to a defined genetic subgroup using our approach of targeted RNA/DNA sequencing. In comparison, 18.7% of cases remained uncharacterized (B other) in contrast to 12.1% B-other cases reported in the St. Jude Total Therapy Study 16.^7^ While this gap highlights the need for broader spectrum genomic profiling in future studies, our approach provides a viable framework that detects most of the WHO5-recognised genetic abnormalities in children with B-ALL in a trial setting.

Here, we developed a three-tier genetic risk model incorporating variables predictive of EFS in a large cohort of children treated uniformly with MRD-directed risk-adapted ICiCLe-ALL-14 protocol. In addition to the high hyperdiploidy and *ETV6::RUNX1*, the SGR group also included *DUX4*-r. Children with *DUX4*-r had a 2-year EFS of 73.3 months (HR-0.73; 95% CI-0.27 to 1.94) in our cohort, and although the presence of DUX4-r was not associated with statistically significant better EFS, possibly due to only 12 cases, *DUX4*-r has been reported to be associated with superior outcome in earlier reports.^7^ *KMT2A*-r are often linked with adverse outcomes in both infants and adults with B-ALL. Our study included a cohort of 1–15-year-olds, and similar to the study where non-infant children with *KMT2A-r* were evaluated,^18^ our cohort of *KMT2A-r* B-ALL demonstrated an intermediate 2-year EFS of 67.5%, thus being grouped into IGR. Interestingly, only the *BCR::ABL1*-like with ABL class-r was associated with inferior outcome (median EFS 31.3 months; HR-2.3; 95% CI-0.68 to 7.55; *p*=0.043) in our cohort and included in HGR, while the JAK-STAT perturbed *BCR::ABL1*-like group (median not reached; HR-1.00; 95% CI-0.37 to 2.65; *p*=0.98) was not associated with significant outcome difference and was included in the IGR category. Although *BCR::ABL1*-like B-ALL is generally associated with inferior prognosis irrespective of age, previous reports have suggested that MRD-directed therapy may be beneficial in abrogating the prognostic impact.^19^ In a retrospective multi-centric study of the pre-tyrosine kinase inhibitor (TKI) era, *BCR::ABL1-*like ABL-class-r had inferior outcomes.^20^ Our cohort of *BCR::ABL1*-like B-ALL has a relatively larger proportion of ABL-class-r cases, which might have allowed the identification of distinct inferior outcomes of this subgroup. Among the patients of *BCR::ABL1-*like ABL-class-r, nine received TKI, despite which their outcome remained poor. The PAX5^alt^ group had the worst 2-year EFS (39.1%, HR-2.8; 95% CI-1.18 to 6.68, *p*=0.0001) in our cohort and is included in the HiR category. Initial studies from the SJRCH/COG group have reported an intermediate prognosis of PAX5^alt^ with MRD-directed therapy,^4,7^ however, based on the dismal EFS in our cohort, we have included the PAX5^alt^ in the HGR category. This finding aligns with the recently published study by Chang and colleagues, where approximately 50% of children with PAX5^alt^ ALL relapsed, highlighting the poor prognosis of this subtype in standard-risk pediatric patients.^21^ We included *IKZF1* deletion, *MEF2D*-r and *TCF3::HLF* in the HGR category, which is in line with previous findings by the SJRCH/COG group.^4,7^

The genetic risk groups proposed here were associated with distinct EFS across ICiCLe risk groups and had an independent association with EFS on multivariate analysis, in addition to ICiCLe-ALL-14 initial risk and PI MRD status. On combining genomic data with PI MRD assessments, we found out that the SGR group was associated with uniformly superior prognosis irrespective of MRD status. This, however, should not be interpreted as a rationale to deescalate therapy as the patients with MRD positive in the SGR category were all treated with high-risk consolidation in this cohort. MRD positive status was associated with inferior outcomes within the IGR and HGR categories, suggesting that a combination of genomic risk group and MRD status might form the most optimum strategy for risk-adapted protocols in children with B-ALL.

We acknowledge the limitation of our study of relatively shorter follow-up and lack of gene expression profiling. Incorporating unbiased sequencing approaches will help further delineate the genetic makeup of the NOS cases and may identify missed cases. However, the stepwise approach, targeted RNA/DNA sequencing strategy, and the genomic risk score described in this manuscript are easily implementable in routine practice and, together with MRD status, may help in refining risk-stratification approaches in children with B-ALL.

In conclusion, we demonstrate the applicability of the WHO5 classification in routine practice and create a general framework for the incorporation of the WHO5 classification in risk-adapted therapy for childhood B-ALL.

## Data Availability

All data produced in the present study are available upon reasonable request to the authors

## Acknowledgements

We would like to express our sincere gratitude to Rakhi Salve, Bhagyashree Satam, Tuhina Srivastava, Harsh Chindarkar and Dr. Shivangi Harankhedkar for their invaluable assistance in entering and managing the patient data for this study. Their contributions were crucial to the successful completion of this work. We also acknowledge Mr Sitaram Ghogale and Mr Nilesh Deshpande for processing samples for flow cytometry testing.

## Authorship and conflict-of-interest statements

SR, NP and GC designed the study, conducted research, analyzed, interpreted the data, and wrote the manuscript. PB, SJ, SC, PT, DS, SG and PS conducted research conducted research and analyzed data. SS, AC, CD, NM, SB and GN recruited patients and analyzed data. Dr Nikhil Patkar is supported by an India Alliance (Wellcome-DBT) Senior Fellowship for Clinicians and Public Health Researchers. Dr Gaurav Chatterjee is supported by an India Alliance (Wellcome-DBT) Intermediate Fellowship for Clinicians and Public Health Researchers. Dr Nikhil Patkar has received research funding from Illumina Inc and ThermoFisher Scientific.

The author(s) declare no competing financial interests.

## Agreement to Share Publication-Related Data and Data Sharing Statement

Dr Nikhil Patkar, Department of Haematopathology, CCE Building, Advanced Centre for Treatment Research and Education in Cancer, Tata Memorial Centre, Kharghar, Maharashtra, India, Pin:410210. Phone: +91-22-27405000

## References

1 Moorman A V., Enshaei A, Murdy D, Joy M, Boer JM, den Boer ML et al. Integration of genetics and MRD to define low risk patients with B-cell precursor acute lymphoblastic leukaemia with intermediate MRD levels at the end of induction. Leukemia 2024; 38. doi:10.1038/S41375-024-02329-0.

2 Moorman A V., Enshaei A, Schwab C, Wade R, Chilton L, Elliott A et al. A novel integrated cytogenetic and genomic classification refines risk stratification in pediatric acute lymphoblastic leukemia. Blood 2014; 124: 1434–1444.

3 Lilljebjörn H, Henningsson R, Hyrenius-Wittsten A, Olsson L, Orsmark-Pietras C, Von Palffy S et al. Identification of ETV6-RUNX1-like and DUX4-rearranged subtypes in paediatric B-cell precursor acute lymphoblastic leukaemia. Nat Commun 2016; 7. doi:10.1038/NCOMMS11790.

4 Gu Z, Churchman ML, Roberts KG, Moore I, Zhou X, Nakitandwe J et al. PAX5-driven Subtypes of B-progenitor Acute Lymphoblastic Leukemia. Nat Genet 2019; 51: 296.

5 Alaggio R, Amador C, Anagnostopoulos I, Attygalle AD, Araujo IB de O, Berti E et al. The 5th edition of the World Health Organization Classification of Haematolymphoid Tumours: Lymphoid Neoplasms. Leukemia. 2022; 36. doi:10.1038/s41375-022-01620-2.

6 Duffield AS, Mullighan CG, Borowitz MJ. International Consensus Classification of acute lymphoblastic leukemia/lymphoma. Virchows Arch 2023; 482: 11–26.

7 Jeha S, Choi J, Roberts KG, Pei D, Coustan-Smith E, Inaba H et al. Clinical Significance of Novel Subtypes of Acute Lymphoblastic Leukemia in the Context of Minimal Residual Disease–Directed Therapy. Blood Cancer Discov 2021; 2: 326–337.

8 Tanasi I, Ba I, Sirvent N, Braun T, Cuccuini W, Ballerini P et al. Efficacy of tyrosine kinase inhibitors in Ph-like acute lymphoblastic leukemia harboring ABL-class rearrangements. Blood 2019; 134: 1351–1355.

9 Gudapati P, Khanka T, Chatterjee G, Ghogale S, Badrinath Y, Deshpande N et al. CD304/neuropilin-1 is a very useful and dependable marker for the measurable residual disease assessment of B-cell precursor acute lymphoblastic leukemia. Cytometry B Clin Cytom 2020; 98: 328–335.

10 Tembhare P, Badrinath Y, Ghogale S, Patkar N, Dhole N, Dalavi P et al. A novel and easy FxCycle^TM^ violet based flow cytometric method for simultaneous assessment of DNA ploidy and six-color immunophenotyping. Cytometry Part A 2016; 89: 281–291.

11 Amare PSK, Jain H, Kabre S, Deshpande Y, Pawar P, Banavali S et al. Cytogenetic Profile in 7209 Indian Patients with <i>de novo</i> Acute Leukemia: A Single Centre Study from India. J Cancer Ther 2016; 07: 530–544.

12 van Dongen JJ, Macintyre EA, Gabert JA, Delabesse E, Rossi V, Saglio G et al. Standardized RT-PCR analysis of fusion gene transcripts from chromosome aberrations in acute leukemia for detection of minimal residual disease. Report of the BIOMED-1 Concerted Action: investigation of minimal residual disease in acute leukemia. Leukemia 1999; 13: 1901–28.

13 Caye A, Beldjord K, Mass-Malo K, Drunat S, Soulier J, Gandemer V et al. Breakpoint-specific multiplex polymerase chain reaction allows the detection of IKZF1 intragenic deletions and minimal residual disease monitoring in B-cell precursor acute lymphoblastic leukemia. Haematologica 2013; 98: 597–601.

14 Das N, Banavali S, Bakhshi S, Trehan A, Radhakrishnan V, Seth R et al. Protocol for ICiCLe-ALL-14 (InPOG-ALL-15-01): a prospective, risk stratified, randomised, multicentre, open label, controlled therapeutic trial for newly diagnosed childhood acute lymphoblastic leukaemia in India. Trials 2022; 23: 1–20.

15 Tembhare PR, Subramanian PG PG, Ghogale S, Chatterjee G, Patkar N V., Gupta A et al. A High-Sensitivity 10-Color Flow Cytometric Minimal Residual Disease Assay in B-Lymphoblastic Leukemia/Lymphoma Can Easily Achieve the Sensitivity of 2-in-106 and Is Superior to Standard Minimal Residual Disease Assay: A Study of 622 Patients. Cytometry B Clin Cytom 2020; 98: 57–67.

16 Alaggio R, Amador C, Anagnostopoulos I, Attygalle AD, Araujo IB de O, Berti E et al. The 5th edition of the World Health Organization Classification of Haematolymphoid Tumours: Lymphoid Neoplasms. Leukemia 2022; 36: 1720–1748.

17 Li JF, Dai YT, Lilljebjörn H, Shen SH, Cui BW, Bai L et al. Transcriptional landscape of B cell precursor acute lymphoblastic leukemia based on an international study of 1,223 cases. Proc Natl Acad Sci U S A 2018; 115: E11711–E11720.

18 Attarbaschi A, Möricke A, Harrison CJ, Mann G, Baruchel A, De Moerloose B et al. Outcomes of Childhood Noninfant Acute Lymphoblastic Leukemia With 11q23/ KMT2A Rearrangements in a Modern Therapy Era: A Retrospective International Study. J Clin Oncol 2023; 41: 1404–1422.

19 Roberts KG, Pei D, Campana D, Payne-Turner D, Li Y, Cheng C et al. Outcomes of children with BCR-ABL1–like acute lymphoblastic leukemia treated with risk-directed therapy based on the levels of minimal residual disease. J Clin Oncol 2014; 32: 3012– 3020.

20 den Boer ML, Cario G, Moorman A V., Boer JM, de Groot-Kruseman HA, Fiocco M et al. Outcomes of paediatric patients with B-cell acute lymphocytic leukaemia with ABL-class fusion in the pre-tyrosine-kinase inhibitor era: a multicentre, retrospective, cohort study. Lancet Haematol 2021; 8: e55–e66.

21 Chang TC, Chen W, Qu C, Cheng Z, Hedges D, Elsayed A et al. Genomic Determinants of Outcome in Acute Lymphoblastic Leukemia. J Clin Oncol 2024. doi:10.1200/JCO.23.02238.

