## Supplementary Data for "Clinical Relevance and Applicability of the 2022 World Health Organization Classification of Childhood B Lymphoblastic Leukemia in the Context of MRD-Directed Therapy"

Newly diagnosed B-ALL (n-533)

Screened for common translocations *ETV6::RUNX1, BCR::ABL1, TCF3::PBX1, KMT2A::AFF1* and *IKZF1* deletion testing (PCR)

Positive for common translocations (n-141)

Negative for common translocations (n-392)

Targeted RNA sequencing (NARASIMHA)

Negative for translocations

Targeted DNA Sequencing (n-124)

Screened for study (n-674)

Excluded from study (n-141)

- Not treated with ICiCLe-ALL-14 protocol (n-127)
- Poor RNA quality (n-14)

**Supplementary Figure 1. Consort diagram indicating testing strategy implemented for B-ALL patient evaluation**

**Methods**

**Risk Stratification:** Patients were stratified according to National Cancer Institute (NCI) risk categories of standard risk (NCI-SR) and high risk (NCI-HR) based on their age and WBC count at diagnosis. Those 10 years or older with a WBC count of 50 × 10^9/L or higher were categorized as NCI-HR. Additional factors were considered to stratify based on ICiCLe-ALL-14 criteria at two time points (day 8- initial) and post-induction (final risk stratification). In the initial risk assessment of the ICiCLe, standard risk included younger B-ALL patients (under 10 years) with low presenting leukocyte counts (less than 50 × 10^9/L) and without high-risk features. Intermediate risk encompassed B-ALL patients lacking high-risk features but who were older and had high presenting leukocyte counts and/or bulky disease. High risk was assigned to B-ALL patients presenting any high-risk feature, including those with high-risk genetics, central nervous system leukemia, or poor prednisolone response on treatment day 8. High-risk features were defined as CNS leukemia, inadequate prednisolone response, insufficient information for risk stratification, including patients with compromised prednisolone response, and those with atypical clinical presentations such as extramedullary disease at unusual sites, and high-risk genetics (*BCR::ABL1*, *KMT2A* rearrangement, iAMP21, hypodiploidy, *TCF3::HLF*). The final ICiCLe-ALL-14 risk stratification also considered post-induction MRD positivity at levels above 0.01%, in addition to the factors considered in the initial ICiCLE risk stratification.^1^

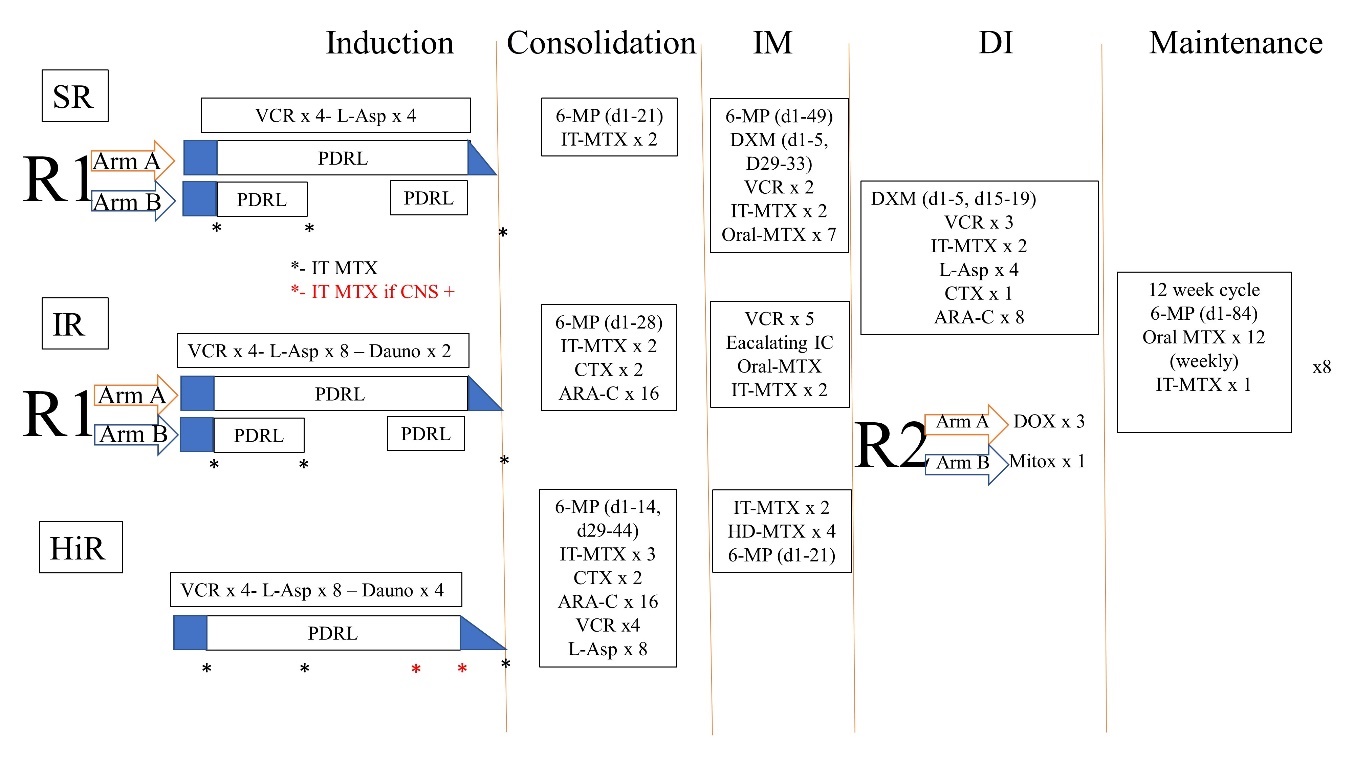

**Supplementary Figure 2. Schematic overview of risk-stratified treatment and randomized interventions for patients diagnosed with B-ALL in the ICiCLe-ALL-14 protocol.** Patients are divided into three risk categories: Standard Risk (SR), Intermediate Risk (IR), and High Risk (HiR). Each risk group undergoes four sequential blocks of intensive therapy—**Induction** (which includes a 7-day prednisolone pre-phase), **Consolidation**, **Interim Maintenance (IM)**, and **Delayed Intensification (DI)**—followed by 24 months of maintenance therapy. Treatment intensity increases with risk category, with the HiR group receiving the most intensive regimen. Younger patients (< 10 years) in the SR and IR categories are randomized in the Induction phase to either the standard continuous prednisolone schedule (R1 Arm A) over 4 weeks with tapering or a shorter, pulsed schedule (R1 Arm B: days 1-14, and 22-28). A second randomization, open to all risk groups, assigns patients in the DI phase to either 3 doses of doxorubicin (R2 Arm A) or a single dose of mitoxantrone (R2 Arm B). Abbreviations: 6-MP, 6-mercaptopurine; ARA-C, cytarabine; CNS+, central nervous system involvement; CTX, cyclophosphamide; Dauno, daunorubicin; DXM, dexamethasone; HD-MTX, high-dose methotrexate; IT-MTX, intrathecal methotrexate; IV, intravenous; L-Asp, E. coli L-asparaginase; MRD, minimal residual disease; MTX, methotrexate; PRDL, prednisolone; VCR, vincristine (Adapted from Das N et al.) ^1^

**RNA sequencing:** Targeted RNA sequencing was performed using the lymphoid module of our in-house assay, NARASIMHA,^2^ with a 25-gene panel as listed in Supplementary Table 1. Fusion detection was carried out on Fastq files through parallel workflows adapted from modification in the nf-core RNA sequencing pipeline.^3^ Briefly, tools such as Arriba v1.2.0, FusionCatcher v1.20, STAR-Fusion v1.8.1, and Pizzly v0.37.3 were employed for fusion detection. After the initial analysis, Fusion-report v2.2.1 was utilized to aggregate and consolidate the identified fusions. Complementary analyses were conducted using FusionInspector v2.2.1, visualization through Arriba v1.2.0. Coverage metrics were determined using BedTools v2.4. A fusion was deemed positive if identified by a minimum of two tools and supported by at least five spanning reads.

**DNA Sequencing:** Children who remained uncharacterized for any specific genetic subgroups after NARASIMHA. underwent DNA sequencing (n=124) using either of two gene panels: 34 (*BCL11B, CCND3, CDKN2A, CDKN2B, CREBBP, CTCF, EZH2, FBXW7, FCGBP, FLT3, IKZF1, IL7R, JAK1, JAK2, JAK3, KDM6A, KMT2D, KRAS, LEF1, MYB, NOTCH1, NRAS, PAX5, PHF6, PTEN, PTPN11, RB1, RPL10, RUNX1, TP53, UNC80, USH2A, USP7, WT1*) or a 7-gene (*PAX5, IKZF1, NRAS, KRAS, TP53, IL7R* and *JAK2*) hotspot panel. Target areas of DNA were enriched by a single-molecule molecular inversion probe (smMIPS) based approach. These smMIPS were designed to target the coding regions and an additional 5 base pairs of intronic flanking of genes known to be involved in ALL pathogenesis. To achieve uniform capture efficiency across targeted regions, the libraries were pooled and then adjusted. For the sequencing process, 600 ng of genomic DNA was processed using the smMIPS panel, followed by exonuclease treatment, PCR amplification, and size selection to create sequencing-ready libraries with dual indices specific to each sample. Sequencing of these libraries was performed using the 300 cycle Illumina MiSeq v2 chemistry system (Illumina, based in San Diego, CA, USA). Sequencing data was analyzed using a customized pipeline that incorporates adapter trimming using ea-utils, read self-assembly using PEAR, bwa (v0.7.12) for mapping to the human genome (build hg19/GRCh37), samtools (v. 0.1.19)& GATK v3.8 for pre-processing the aligned file. For the identification of sequence variants, we adhered to the GATK best practices framework. Variant calling was done using Varscan (ver2.3), MuTect2.0, and Platypus v0.8.1. The identified variants were subsequently annotated, referencing several population frequency databases using Annovar. These include the 1000 Genomes Project (covering diverse populations such as African, Admixed American, East Asian, Finnish, Non-Finnish European, and South Asian), the Exome Aggregation Consortium datasets, NHLBI-ESP (encompassing 6500 genomes), and the COSMIC database (versions 80 and 83). Variants were filtered, focusing on exonic and spicing variants. The variants that were detected in population frequency of >0.01 were removed and the remaining variants were subjected to in-silico prediction tools (SIFT, PolyPhen2, CADD, PROVEAN, MutationTaster, MutationAssessor, M-CAP, FATHMM, LRT, DANN) for determination of their pathogenicity. Variants that were called by atleast 2 variant callers and determined pathogenic or likely pathogenic were reported.

| **Gene** | **RefSeq ID** | **Region** | **Gene** | **RefSeq ID** | **Region** |
| --- | --- | --- | --- | --- | --- |
| *ABL1* | NM005157 | Exons 1-5 | *KMT2A* | NM005933 | Exons 2-35 |
| *ABL2* | NM005158 | Exons 2-8 | *MEF2D* | NM005920 | Exons 5-8 |
| *CRLF1* | NM022147 | Exon 1 | *NTRK3* | NM001007156 | Exon 15 |
| *CRLF2* | NM022148 | Exons 1-6 | *NUP98* | NM016320 | Exons 12-17 |
| *CSF1R* | NM005211 | Exons 9-14 | *NUP214* | NM005085 | Exons 17-19 |
| *DUX4* | NM001293798 | Exon 1 | *PDGFRA* | NM006206 | Exons 9-12,14 |
| *EBF1* | NM024007 | Exons 10-15 | *PDGFRB* | NM002609 | Exons 8-9,13 |
| *EPOR* | NM000121 | Exons 7-8 | *P2RY8* | NM178129 | Exon 1 |
| *ETV6* | NM001987 | Exons 1-6 | *PAX5* | NM016734 | Exons 1,4-8 |
| *FGFR1* | NM023110 | Exons 2-12,17 | *RUNX1* | NM001754 | Exons 2-9 |
| *IKZF1* | NM006060 | Exons 1-3,7,8 | *TCF3* | NM003200 | Exons 11-18 |
| *IL2RB* | NM000878 | Exon 10 | *ZNF384* | NM001039920 | Exons 2,3,7 |
| *JAK2* | NM004972 | Exons 6-13, 15,20 |  |  |  |

**Supplementary Table 1. Panel of genes tested using the targeted RNA sequencing approach (NARASIMHA)**

**Results**

**MRD Characteristics of Genetic Subtypes:** More than half of the cases in the *BCR::ABL1* (p=0.02), *BCR::ABL1*-like (p<0.0001), *TCF3::HLF* (p=0.19), and *ZNF384* rearrangement (p=0.51) groups exhibited positive PI MRD. Significantly, in the *TCF3::HLF*, *BCR::ABL1* like, *ZNF384* rearrangement, *BCR::ABL1* and hypodiploidy categories, a significant proportion of patients had MRD>1% comprising of 50, 39.13, 33.33, 24, and 16.67% cases respectively. The proportion of patients having PI MRD<0.01, 0.01-1%, 1-5% and >5% MRD in each genetic categories is shown in Supplementary Figure 2.

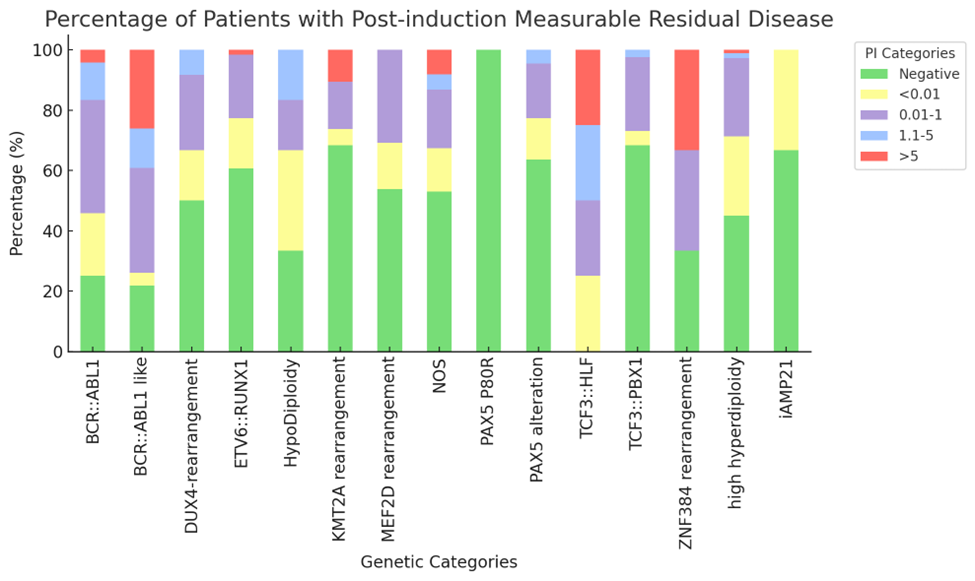

**Supplementary Figure 3. Post induction MRD levels in patients of genetic sub-categories**

|  | Percentage of IKZF1 deletion |
| --- | --- |
| BCR::ABL1 | 56.5217 |
| BCR::ABL1 like | 55.5556 |
| DUX4-r | 16.6667 |
| KMT2A-r | 15.3846 |
| NOS | 12 |
| ETV6::RUNX1 | 1.63934 |
| high hyperdiploidy | 1.28205 |

**Supplementary Figure 4. *IKZF1* deletion frequencies across genetic sub-groups.**

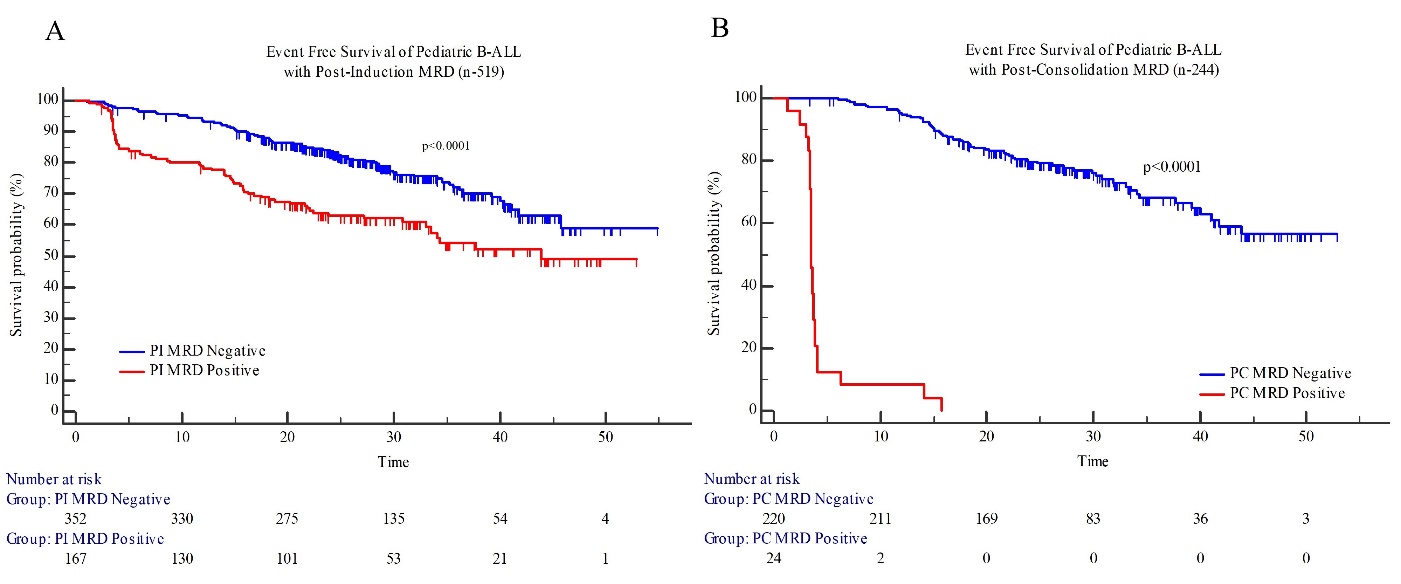

**Supplementary Figure 5. Event-free survival of pediatric B-ALL patients with post-induction and post-consolidation measurable residual disease.**

**Impact of** **ICiCLe Risk stratification on Patient Outcomes:** The 2-year EFS rates based on ICiCLe initial risk categorization were 86.2% for standard risk (SR), 70.6% for intermediate risk (IR), and 67.0% for HiR, respectively (*p*<0.0001; supplementary Figure 5A). Similarly, the 2-year OS based on ICiCLe initial risk categorization were 92.8% for SR, 76.7% for IR and 83.2% for HiR (*p*<0.0001). For the ICiCLe final risk categorization, the 2-year EFS rates were 89.9% for SR, 79.0% for IR, and 70.9% for HiR (p<0.0001; supplementary Figure 5B) and the corresponding 2-years OS were 94.4% for SR, 83.3% for IR and 81.6% for HiR respectively (*p*=0.242).

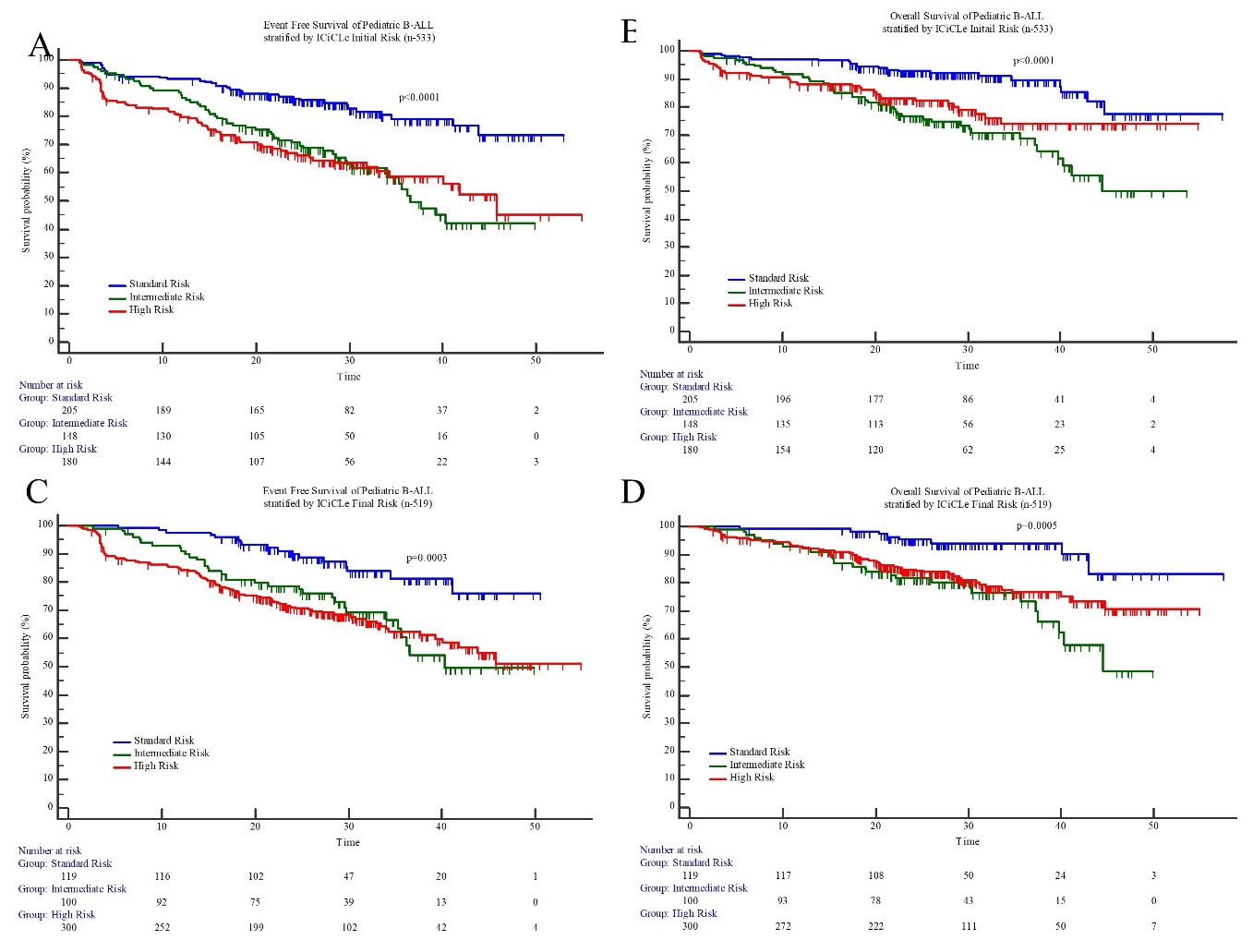

**Supplementary Figure 6. Event free and Overall survival of pediatric B-ALL patients stratified by ICiCLe initial and final risk.**

| **Event free survival** | | | | **Overall survival** | | | |
| --- | --- | --- | --- | --- | --- | --- | --- |
| **Variable** | **HR** | **95% CI** | ***p*-value** | **Variable** | **HR** | **95% CI** | ***p*-value** |
| ICiCLe Initial Risk | 1.29 | 1.05 to 1.57 | 0.0154 | ICiCLe Initial Risk | 1.21 | 0.93 to 1.57 | 0.148 |
| Post induction MRD | 1.98 | 1.42 to 2.74 | <0.0001 | Post induction MRD | 1.57 | 1.03 to 2.38 | 0.037 |
| Genetic Risk | 1.99 | 1.62 to 2.45 | <0.0001 | Genetic Risk | 1.67 | 1.29 to 2.17 | 0.0001 |

**Supplementary Table 2. Multivariate regression analysis of factors influencing event-free survival and overall survival. HR- Hazard ratio, CI- Confidence Interval**

**References:**

1	Das N, Banavali S, Bakhshi S, Trehan A, Radhakrishnan V, Seth R *et al.* Protocol for ICiCLe-ALL-14 (InPOG-ALL-15-01): a prospective, risk stratified, randomised, multicentre, open label, controlled therapeutic trial for newly diagnosed childhood acute lymphoblastic leukaemia in India. *Trials* 2022; **23**: 1–20.

2	Patkar N, Bhanshe P, Rajpal S, Joshi S, Chaudhary S, Chatterjee G *et al.* NARASIMHA: Novel Assay based on Targeted RNA Sequencing to Identify ChiMeric Gene Fusions in Hematological Malignancies. *Blood Cancer Journal 2020 10:5* 2020; **10**: 1–4.

3	Ewels PA, Peltzer A, Fillinger S, Patel H, Alneberg J, Wilm A *et al.* The nf-core framework for community-curated bioinformatics pipelines. *Nature Biotechnology 2020 38:3* 2020; **38**: 276–278.
